# DyeVert™ PLUS EZ system for Preventing Contrast-Induced Acute Kidney Injury in Patients Undergoing Diagnostic Coronary Angiography and/or Percutaneous Coronary Intervention: A UK-Based Cost-Utility Analysis

**DOI:** 10.1101/19008185

**Authors:** Mehdi Javanbakht, Mohsen Rezaei Hemami, Atefeh Mashayekhi, Michael Branagan-Harris

## Abstract

**Background:** Contrast-induced acute kidney injury (CI-AKI) is a complication commonly associated with invasive angiographic procedures and is considered the leading cause of hospital-acquired acute kidney injury. CI-AKI can lead to a prolonged hospital stay, with a substantial economic impact, and increased mortality. The DyeVert™ PLUS EZ system (FDA approved and CE marked) is a device that has been developed to divert a portion of the theoretical injected contrast media volume (CMV), reducing the overall injected contrast media and aortic reflux and potentially improving long-term health outcomes.

**Objectives:** To assess the long-term costs and health outcomes associated with the introduction of the DyeVert™ PLUS EZ system into the health care service for the prevention of CI-AKI in a cohort of patients with chronic kidney disease (CKD) stage 3-4 undergoing Diagnostic Coronary Angiography (DAG) and/or Percutaneous Coronary intervention (PCI), compared with current practice.

**Methods:** A de novo economic model was developed based on the current pathway of managing patients undergoing DAG and/or PCI and on evidence related to the clinical effectiveness of DyeVert™, in terms of its impact on relevant clinical outcomes and health service resource use. Clinical data used to populate the model were derived from the literature or were based on assumptions informed by expert clinical input. Costs included in the model were obtained from the literature and UK-based routine sources. Probabilistic distributions were assigned to the majority of model parameters so that a probabilistic analysis could be undertaken, while deterministic sensitivity analyses were also carried out to explore the impact of key parameter variation on the model results.

**Results:** Base-case results indicate that the intervention leads to cost savings (- £3,878) and improved effectiveness (+ 0.02 QALYs) over the patient’s lifetime, compared with current practice. Output from the probabilistic analysis supports the high likelihood of the intervention being cost-effective across presented willingness-to-pay (WTP) thresholds. The overall long-term cost saving for the NHS associated with introduction of the intervention for each cohort of patients is over £175 million. The cost savings are mainly driven by lower risk of subsequent diseases and associated costs

**Conclusions:** Introduction of the DyeVert™ PLUS EZ system has the potential to reduce costs for the health care service and lead to improved clinical outcomes for patients with CKD stage 3-4 undergoing angiographic procedures.

**Key Points for Decision Makers:**

- An economic model has been developed to consider the cost-effectiveness of the DyeVert™ PLUS EZ system for use amongst patients undergoing angiographic procedures.
- Results of the economic analysis indicate that the DyeVert™ PLUS EZ system is highly likely to be cost saving and result in improved patient outcomes.

## 1. Introduction

One common complication associated with angiographic procedures, and attributed to radiocontrast media, is the contrast-induced acute kidney injury (CI-AKI) (1–3). CI-AKI is defined as the impairment of renal function, measured as either a 25% increase in serum creatinine (SCr) from baseline or a 0.5 mg/dL (44 µmol/L) increase in absolute SCr value within 48-72 hours after intravenous contrast administration.

The development of CI-AKI can lead to prolonged hospital stay, additional financial burden, and increased mortality (4, 5). In particular, the economic impact of CI-AKI in the UK is significant. It is estimated that in 2010-2011 there were 977,116 excess bed days attributable to AKI in the UK, with an associated cost of £304 million (4). CI-AKI is considered the leading cause of hospital-acquired AKI and is responsible for one third of AKI cases (6). This condition is closely related to angiographic procedures, where the incidence of CI-AKI in patients undergoing angiographic procedures ranges between 1-2% for patients without prior chronic kidney disease (CKD), and 30% for patients with a combination of risk factors (7–9).

Since there is no definitive treatment available for CI-AKI, the focus has been on the prevention of the condition. The European Society of Cardiology (ESC) published updated guidelines on the prevention of CI-AKI (10), providing a framework for the use of evidence-based preventative strategies. Among these strategies, they recommend the identification of patients at risk of CI-AKI, appropriate periprocedural hydration, and minimising contrast volume in at-risk patients. Previous studies have shown that the CI-AKI is associated with increased risk of death, myocardial infarction, bleeding, and recurrent renal injury after discharge (11, 12).

The DyeVert™ PLUS EZ system (FDA approved and CE marked) is a device that adjusts contrast media (CM) during manual injection. This is achieved by diverting a portion of the theoretical injected contrast media volume (CMV), reducing the overall injected contrast media and reducing aortic reflux (13). Reduction in CMV has been shown to be non-linearly associated with decreased risk of CI-AKI, which can be linked to a reduction in short- and long-term costs and consequences (14). This economic analysis aims to estimate the cumulative difference in costs and effectiveness in patients undergoing Diagnostic Coronary Angiography (DAG) and/or Percutaneous Coronary intervention (PCI) with the use of DyeVert™ PLUS compared to the current standard of care.

## 2. Methods

A de novo economic model was developed reflecting the current management pathway of patients undergoing DAG and/or PCI. The model was built upon evidence related to the clinical effectiveness of DyeVert™ PLUS, measured as the reduction in contrast media (CM) and subsequent incidence of CI-AKI, as well as economic evidence related to associated NHS resource use. For each treatment arm, costs and outcomes were aggregated on the basis of a series of decisions and events. The structure of the model remained unchanged between the two treatment options.

The model was based on a hypothetical cohort of patients with CKD stage 3-4 undergoing DAG and/or PCI. As per NICE requirements, and in order to fully capture the differences in costs and benefits between the DyeVert™ PLUS EZ system and the comparator, a lifetime time horizon was used in the base-case analysis (15). The recommended discount rate in the UK (3.5% per annum) was applied for both costs and benefits (16). The model considered all costs from the UK NHS and personal social services perspective, and was developed in Microsoft Excel.

### 2.1 Model structure

The model structure comprises a decision tree followed by a Markov model with six health states. This model has a lifetime time-horizon with costs and benefits estimated in the decision tree for the first three months, and in the Markov model for the remainder of the patient’s lifetime. The Markov model has a cycle length of three months. Figure 1 presents an illustrative representation of the model structure. The model was used to simulate the management of patients undergoing DAG and/or PCI with reduced kidney function (i.e. eGFR 15 - 60 ml/min/1.73m2). In each strategy, patients may or may not experience a CI-AKI requiring further treatment. Patients may then either remain in state ‘CKD stage 3-4’ (or state ‘CKD stage 3-4 (AKI history)’ if they previously had CI-AKI) or progress according to the natural progression of CKD to state ‘CKD stage 5’. Patients whose renal insufficiency is not severe (i.e. ‘CKD stage 3-4’) can experience a recurrent AKI or a myocardial infarction (MI) at any point. Patients who enter the ‘CKD stage 5’ are assumed to either remain in this state or die, as they will be receiving dialysis and treatments to prevent an MI. Patients in each model state will incur associated costs and quality-adjusted life-years (QALYs).

**Figure 1.**
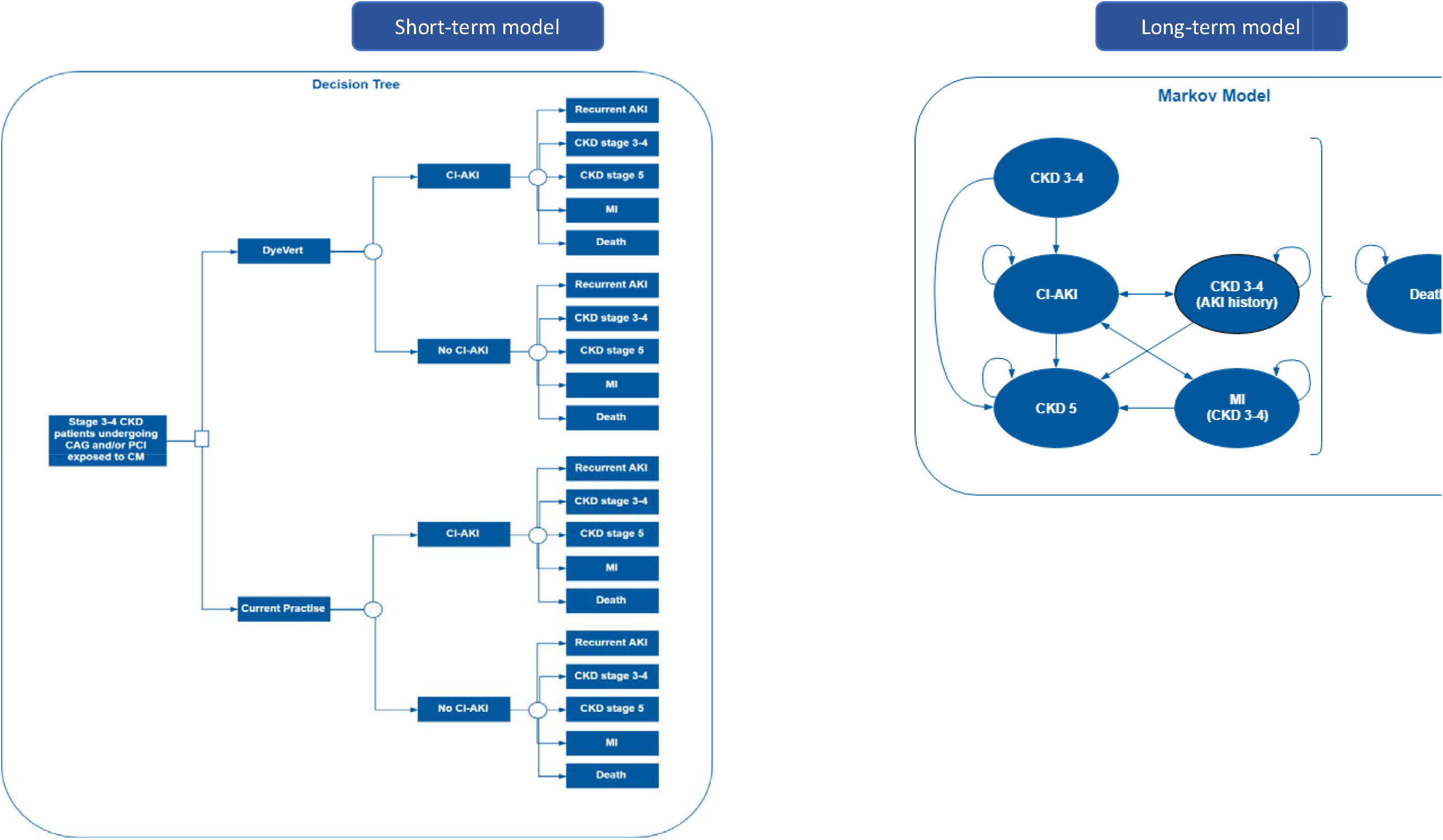
Model Structure.

Simulated patients are at risk of death from all causes during any given cycle period. Risk of death is conditional on CKD stage, history of AKI and/or MI, and age. The all-cause mortality rates were derived from general population mortality statistics reported in national life tables (17) and were adjusted to reflect the extra mortality associated with CI-AKI and renal insufficiency.

Cost-utility analysis of DyeVert™ PLUS EZ system for Preventing Contrast-Induced Acute Kidney Injury

### 2.2 Model inputs

All inputs used to populate the economic model are described in the following section and are presented in Table 2. Patients at model entry was those undergoing DAG and/or PCI with some kidney function impairment (CKD stage 3-4). The base-case population was 72 years old (14). The number of patients undergoing DAG and/or PCI was derived from NHS reference costs activity of percutaneous coronary angioplasty (HRG code EY40-EY41) and cardiac catheterisation (HRG code EY42-EY43) (18). It was assumed, based on Gurm et al. (14) that 26% of these procedures would be DAG combined with PCI.

**Table 1.** Effectiveness of DyeVert™ PLUS.

| Effectiveness | Value (SD) | Source |
| --- | --- | --- |
| % Contrast volume reduction per procedure (DyeVert™) | 40.1% (8.8) | (14) |
| % Reduction in Risk of CI-AKI associated with 40.1% reduction in contrast media for patients with eGFR ≤ 60 ml/min/1.73m<sup>2</sup> | 15.1% | (19) |

**Table 2.**
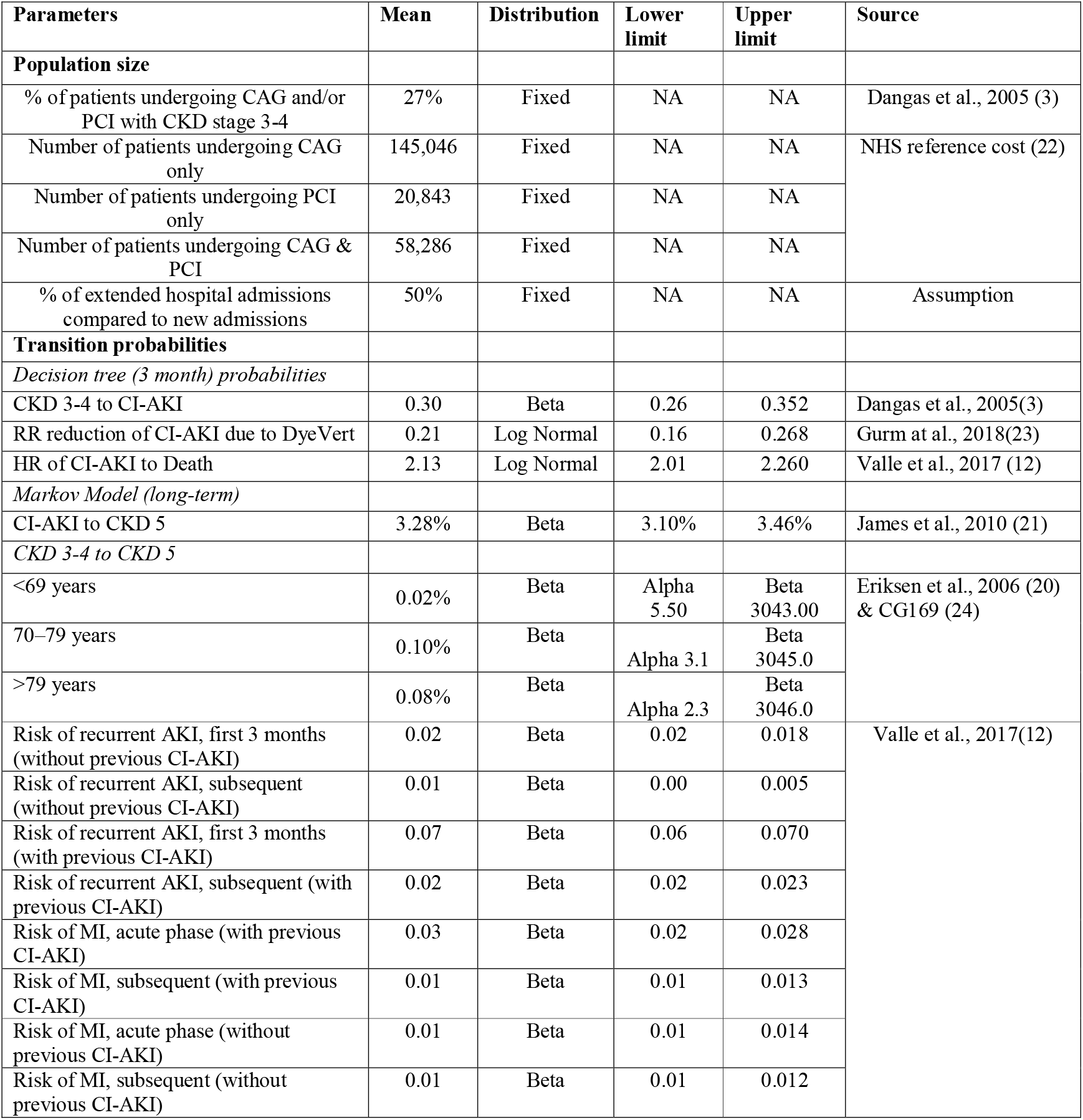
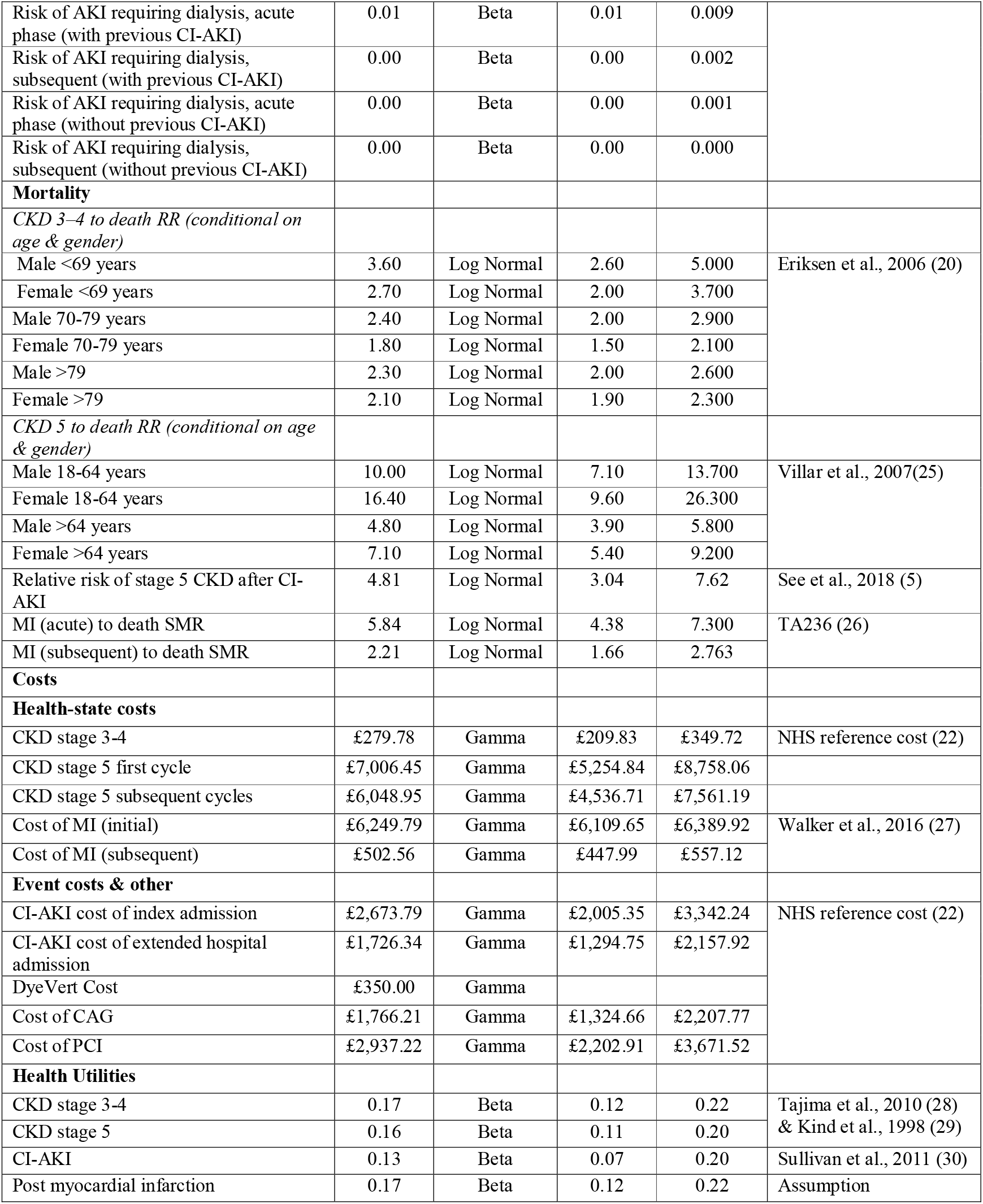
Main inputs used in the model.

| Parameters | Mean | Distribution | Lower limit | Upper limit | Source |
| --- | --- | --- | --- | --- | --- |
| **Population size** |  |  |  |  |  |
| % of patients undergoing CAG and/or PCI with CKD stage 3-4 | 27% | Fixed | NA | NA | Dangas et al., 2005 (3) |
| Number of patients undergoing CAG only | 145,046 | Fixed | NA | NA | NHS reference cost (22) |
| Number of patients undergoing PCI only | 20,843 | Fixed | NA | NA |  |
| Number of patients undergoing CAG & PCI | 58,286 | Fixed | NA | NA |  |
| % of extended hospital admissions compared to new admissions | 50% | Fixed | NA | NA | Assumption |
| **Transition probabilities** |  |  |  |  |  |
| *Decision tree (3 month) probabilities* |  |  |  |  |  |
| CKD 3-4 to CI-AKI | 0.30 | Beta | 0.26 | 0.352 | Dangas et al., 2005(3) |
| RR reduction of CI-AKI due to DyeVert | 0.21 | Log Normal | 0.16 | 0.268 | Gurm et al., 2018(23) |
| HR of CI-AKI to Death | 2.13 | Log Normal | 2.01 | 2.260 | Valle et al., 2017 (12) |
| *Markov Model (long-term)* |  |  |  |  |  |
| CI-AKI to CKD 5 | 3.28% | Beta | 3.10% | 3.46% | James et al., 2010 (21) |
| *CKD 3-4 to CKD 5* |  |  |  |  |  |
| <69 years | 0.02% | Beta | Alpha 5.50 | Beta 3043.00 | Eriksen et al., 2006 (20) & CG169 (24) |
| 70–79 years | 0.10% | Beta | Alpha 3.1 | Beta 3045.0 |  |
| >79 years | 0.08% | Beta | Alpha 2.3 | Beta 3046.0 |  |
| Risk of recurrent AKI, first 3 months (without previous CI-AKI) | 0.02 | Beta | 0.02 | 0.018 | Valle et al., 2017(12) |
| Risk of recurrent AKI, subsequent (without previous CI-AKI) | 0.01 | Beta | 0.00 | 0.005 |  |
| Risk of recurrent AKI, first 3 months (with previous CI-AKI) | 0.07 | Beta | 0.06 | 0.070 |  |
| Risk of recurrent AKI, subsequent (with previous CI-AKI) | 0.02 | Beta | 0.02 | 0.023 |  |
| Risk of MI, acute phase (with previous CI-AKI) | 0.03 | Beta | 0.02 | 0.028 |  |
| Risk of MI, subsequent (with previous CI-AKI) | 0.01 | Beta | 0.01 | 0.013 |  |
| Risk of MI, acute phase (without previous CI-AKI) | 0.01 | Beta | 0.01 | 0.014 |  |
| Risk of MI, subsequent (without previous CI-AKI) | 0.01 | Beta | 0.01 | 0.012 |  |
| Risk of AKI requiring dialysis, acute phase (with previous CI-AKI) | 0.01 | Beta | 0.01 | 0.009 |  |
| Risk of AKI requiring dialysis, subsequent (with previous CI-AKI) | 0.00 | Beta | 0.00 | 0.002 |  |
| Risk of AKI requiring dialysis, acute phase (without previous CI-AKI) | 0.00 | Beta | 0.00 | 0.001 |  |
| Risk of AKI requiring dialysis, subsequent (without previous CI-AKI) | 0.00 | Beta | 0.00 | 0.000 |  |
| **Mortality** |  |  |  |  |  |
| *CKD 3–4 to death RR (conditional on age & gender)* |  |  |  |  |  |
| Male <69 years | 3.60 | Log Normal | 2.60 | 5.000 | Eriksen et al., 2006 (20) |
| Female <69 years | 2.70 | Log Normal | 2.00 | 3.700 |  |
| Male 70-79 years | 2.40 | Log Normal | 2.00 | 2.900 |  |
| Female 70-79 years | 1.80 | Log Normal | 1.50 | 2.100 |  |
| Male >79 | 2.30 | Log Normal | 2.00 | 2.600 |  |
| Female >79 | 2.10 | Log Normal | 1.90 | 2.300 |  |
| *CKD 5 to death RR (conditional on age & gender)* |  |  |  |  |  |
| Male 18-64 years | 10.00 | Log Normal | 7.10 | 13.700 | Villar et al., 2007(25) |
| Female 18-64 years | 16.40 | Log Normal | 9.60 | 26.300 |  |
| Male >64 years | 4.80 | Log Normal | 3.90 | 5.800 |  |
| Female >64 years | 7.10 | Log Normal | 5.40 | 9.200 |  |
| Relative risk of stage 5 CKD after CI-AKI | 4.81 | Log Normal | 3.04 | 7.62 | See et al., 2018 (5) |
| MI (acute) to death SMR | 5.84 | Log Normal | 4.38 | 7.300 | TA236 (26) |
| MI (subsequent) to death SMR | 2.21 | Log Normal | 1.66 | 2.763 |  |
| **Costs** |  |  |  |  |  |
| **Health-state costs** |  |  |  |  |  |
| CKD stage 3-4 | £279.78 | Gamma | £209.83 | £349.72 | NHS reference cost (22) |
| CKD stage 5 first cycle | £7,006.45 | Gamma | £5,254.84 | £8,758.06 |  |
| CKD stage 5 subsequent cycles | £6,048.95 | Gamma | £4,536.71 | £7,561.19 |  |
| Cost of MI (initial) | £6,249.79 | Gamma | £6,109.65 | £6,389.92 | Walker et al., 2016 (27) |
| Cost of MI (subsequent) | £502.56 | Gamma | £447.99 | £557.12 |  |
| **Event costs & other** |  |  |  |  |  |
| CI-AKI cost of index admission | £2,673.79 | Gamma | £2,005.35 | £3,342.24 | NHS reference cost (22) |
| CI-AKI cost of extended hospital admission | £1,726.34 | Gamma | £1,294.75 | £2,157.92 |  |
| DyeVert Cost | £350.00 | Gamma |  |  |  |
| Cost of CAG | £1,766.21 | Gamma | £1,324.66 | £2,207.77 |  |
| Cost of PCI | £2,937.22 | Gamma | £2,202.91 | £3,671.52 |  |
| **Health Utilities** |  |  |  |  |  |
| CKD stage 3-4 | 0.17 | Beta | 0.12 | 0.22 | Tajima et al., 2010 (28) & Kind et al., 1998 (29) |
| CKD stage 5 | 0.16 | Beta | 0.11 | 0.20 |  |
| CI-AKI | 0.13 | Beta | 0.07 | 0.20 | Sullivan et al., 2011 (30) |
| Post myocardial infarction | 0.17 | Beta | 0.12 | 0.22 | Assumption |

### Clinical effectiveness

Evidence has shown that when DyeVert™ PLUS is used in patients undergoing DAG and/or PCI the contrast media volume is significantly reduced (14). To estimate the reduction in risk of CI-AKI after using DyeVert™, we analyzed the CI-AKI rate based on the 0.5 serum creatinine increase definition used in the development of the Mehran (9) risk score and ran the Mehran risk score on the population to arrive at a projected risk rate for the population. The projected rate was 14% and the actual rate was 11%. Therefore, the estimated risk reduction was estimated to be 21.4%. The estimated risk reduction was used to adjust the risk of CI-AKI in the intervention arm for the base-case analysis. Alternatively, the risk reduction was estimated using the data from Gurm et al., 2016 and Gurm et al., 2018 (14, 19) (Table 1 below).

### Transition probabilities

The incidence of CI-AKI in patients with CKD undergoing PCI was based on a cohort study of 1,473 patients and was estimated to be 30% (3, 9). The baseline transition probability associated with the progression of patients from CKD stage 3-4 to CKD stage 5 for different age groups, was based on a ten-year cumulative incidence rate in a cohort study of 3,047 patients (20). The probability of transitioning to Stage 5 CKD following a CI-AKI after the first cycle (first 3 months) was obtained from clinical guidelines from CG169 and James et al., 2010 (21) and from the study by Valle et al., 2017 (12). The probability of MI for patients with a history of CI-AKI was taken from Valle et al., 2017 (12). For patients who had not experienced an AKI throughout the model, the probability of MI was 1.42% and 0.688% in the first 3 months and the subsequent cycles respectively (12). For patients who had experienced CI-AKI in the previous cycles, the equivalent probabilities were 2.581% and 1.291% respectively (12).

The probability of recurrent AKI was taken from the same study as the probability of MI (12). Based on the cumulative incidence in the 3rd month and the first year of the study follow-up, we estimated the probability of recurrent AKI in the first 3 months after the DAG and/or PCI, and for subsequent 3-month cycles. For patients who had not experienced a CI-AKI after the procedure, the probability of recurrent AKI was 1.78% and 0.911% in the first 3 months and the subsequent cycles respectively. For patients who had experienced CI-AKI during the procedure, the equivalent probabilities were 6.83% and 2.431% respectively.

### Probability of death

Standardised mortality ratios for each health state included in the model were applied to the relevant age-dependent mortality rates and are shown in Table 2 below. These mortality ratios were derived from the literature.

### Costs

#### Resource consequences

Unit costs for all resource use estimates were extracted from the literature or obtained through other relevant sources such as NHS reference costs (22), Personal Social Services Research Unit (31), British national Formulary and manufacturer price list (32). Costs were measured in Sterling (£) for the year 2018 and were discounted at 3.5% per annum, where appropriate. All costs included in the model are shown in Table 2.

The choice of cost items was mainly informed by a NICE clinical guidelines model developed for the evaluation of prevention strategies of CI-AKI using different hydration methods (33). However, updated unit costs and dosages of drugs were extracted from the literature. In the instances where unit costs of relevant outcomes were not available, the cost of items used in the NICE guidelines model (CG169 (24)) were inflated to reflect current prices. This was done by applying an inflation index provided by the National bank of England (34).

#### Intervention cost

The cost of DyeVert™ PLUS technology was estimated to be £350, including the cost of the smart syringe and module. The company has indicated that free training for using the technology will be provided to clinical staff, and a smart monitor will be provided free of charge. Therefore, these costs were not taken into account.

#### CI –AKI costs

For the estimation of CI-AKI event costs, two different methods were used in the model: (1) For patients who have to be re-admitted to the hospital due to CI-AKI after a DAG and/or PCI procedure, the cost of an index admission due to AKI was used. (2) For patients admitted for DAG and/or PCI who have a prolonged length of stay due to CI-AKI, the cost of these additional bed days was considered.

#### Health state costs

Patients in CKD stage 3-4 are expected to incur costs associated with consultations with the nephrologist (33), combined with lab resource costs and an assumed 5-minute phlebotomist time to measure the patient’s eGFR. Additional costs would include 9% of patients requiring Epoetin-alfa to treat anaemia, as recommended by the clinical guidelines for anaemia treatment in patients with CKD (33). Epoetin-alfa dosage was estimated for a 77kg individual on average, according to ONS. To take into consideration the patients who require diuretics, an assumption was made based on the guidelines model (33) that about a quarter of patients (26%) in the CKD 3-4 stage were in CKD stage 4, 60% of whom would be on a 40mg daily dose of Furosemide (24). The total cost of CKD stage 3-4 per cycle was estimated to be £269.50.

Patients in stage 5 CKD will incur the drug costs. However, patients in this stage will also incur costs associated with Renal Replacement Therapy (RRT) or Conservative Management. In this stage, patients are expected to incur costs such as RRT procedures, anaemia management, specialist appointments, EGFR measurements and diuretics. In the first cycle that a patient enters the CKD stage 5 state, it is assumed that the intensity of treatment will be increased compared to later stages, as costs of initiating treatment are captured. It was assumed, based on the NICE guidelines model (24), that 90% of patients would receive RRT in this model state. For the estimation of RRT costs, a pooled average was taken from NHS reference costs 2017-18 accounting for national usage of different treatment modalities, such as haemodialysis and filtration (22). Patients entering the CKD stage 5 will receive an access procedure, which will then allow for permanent access for RRT. In subsequent cycles, it was assumed that there would be no further access procedure-related costs. Drugs and check-ups are also required and are more frequent in ‘CKD stage 5’. It was assumed that all patients in this stage would have an eGFR more frequently (on a weekly basis), and two nephrologist appointments per three months. In this state, Epoetin was assumed to be administered to 33% of the patients in the same dosage as for patients in CKD 3-4 stages (33). Patients on conservative management (10%) will be receiving monthly home visits by a specialist nurse as well as telephone calls on a weekly basis. It was assumed (33) that diuretics would be used by 90% of the patients with a double dosage compared to CKD 3-4 stages (80mg). The cost of AMI was taken from Walker et al., 2016 (27). In this study, the cost of AMI for the 1^st^ year was estimated to be £6,869 and £1,780 for subsequent years. These costs were inflated to represent prices in 2018.

#### Utilities

Utilities for CKD stages were obtained from a Japanese study (28) used in both the NICE guidelines (CG169) (24, 33) and an NHS report on kidney care (35). An adjustment for the UK population was made by multiplying the values with the general UK population average utility values for people aged 65 to 75. For the stage of CI-AKI, utility values were taken from Sullivan et al., 2011 (30) who reported in a catalogue of UK EQ-5D derived utilities a 0.525 (n=194) utility value associated with kidney injury (‘renal failure’). For the MI state, the same utility as in CKD 3-4 was assumed.

## 3. Analysis

A Monte Carlo simulation was conducted (10,000 iterations) to derive cumulative estimates of costs and effects for each strategy in the model. Probabilistic distributions were assigned to the majority of model parameters (Table 2) so that a probabilistic analysis could be undertaken, while deterministic sensitivity analyses were also carried out to explore the impact of key parameter variation on the model results. Total cost savings for the entire cohort of patients receiving the intervention in the UK over a lifetime time horizon were also calculated.

## 4. Results

This section presents the results of the economic analysis; base-case results are presented first followed by results of the sensitivity analysis.

### Base-case analysis

Results of the base-case analysis presented in Table 3 indicate that the introduction of DyeVert™ PLUS EZ system leads to cost savings of £3,878 per patient over a lifetime time horizon. Additionally, the intervention leads to improved effectiveness over the patient’s lifetime (+0.02). Therefore, DyeVert™ PLUS is considered a dominant strategy (less costly and more effective) compared to current practice.

**Table 3.** Base-case probabilistic results.

| Base-case probabilistic results | Current Practice | DyeVert™ PLUS |
| --- | --- | --- |
| Cost (£) | £24,507 | 20,630 |
| Incremental cost (£) | -£3,878 |  |
| QALYs | 4.54 | 4.56 |
| Incremental QALYs | +0.02 |  |
| ICER (£) ($\Delta\text{Cost}/\Delta\text{QALYs}$) | DyeVert™ PLUS Dominant | |
| Probability of being cost-effective at £20,000 WTP threshold | 100% |  |
| Probability of being cost saving | 100% |  |

The scatter plot produced from the probabilistic analysis (Figure 2) shows that the vast majority of points from the 10,000 iterations of the model are in the South-East quadrant of the cost-effectiveness plane (less costly and more effective), while all simulations indicate that the intervention is less costly than the comparator. Additionally, the cost-effectiveness acceptability curve (CEAC) shown in Figure 3 (displaying the probability that the intervention is cost-effective across a range of willingness-to-pay (WTP) thresholds) indicates that the DyeVert™ PLUS EZ system has a 100% probability of being cost-effective across all WTP thresholds presented. The overall long-term cost savings for the NHS for cohort of patients is over £175 million (Table 4). Total cost savings are calculated based on the total number of patients receiving the intervention in the UK over a lifetime time horizon. The cost savings are mainly driven by lower risk of subsequent diseases and associated costs.

**Table 4.**
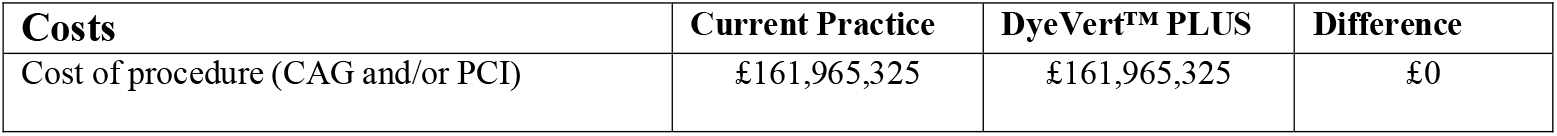
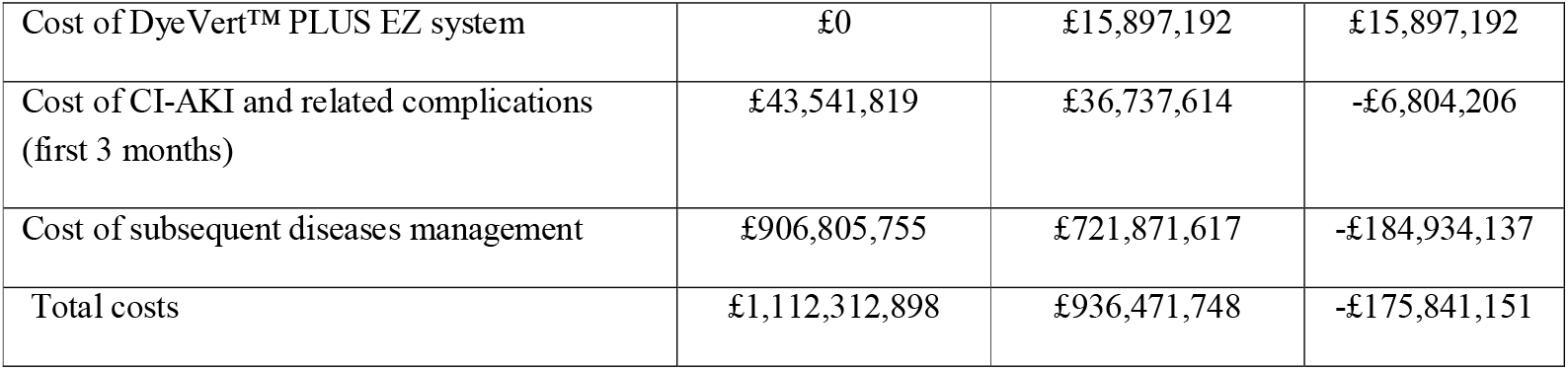
Total long-term costs results (per cohort of patients)

| Costs | Current Practice | DyeVert™ PLUS | Difference |
| --- | --- | --- | --- |
| Cost of procedure (CAG and/or PCI) | £161,965,325 | £161,965,325 | £0 |
| Cost of DyeVert™ PLUS EZ system | £0 | £15,897,192 | £15,897,192 |
| Cost of CI-AKI and related complications (first 3 months) | £43,541,819 | £36,737,614 | -£6,804,206 |
| Cost of subsequent diseases management | £906,805,755 | £721,871,617 | -£184,934,137 |
| Total costs | £1,112,312,898 | £936,471,748 | -£175,841,151 |

**Figure 2.**
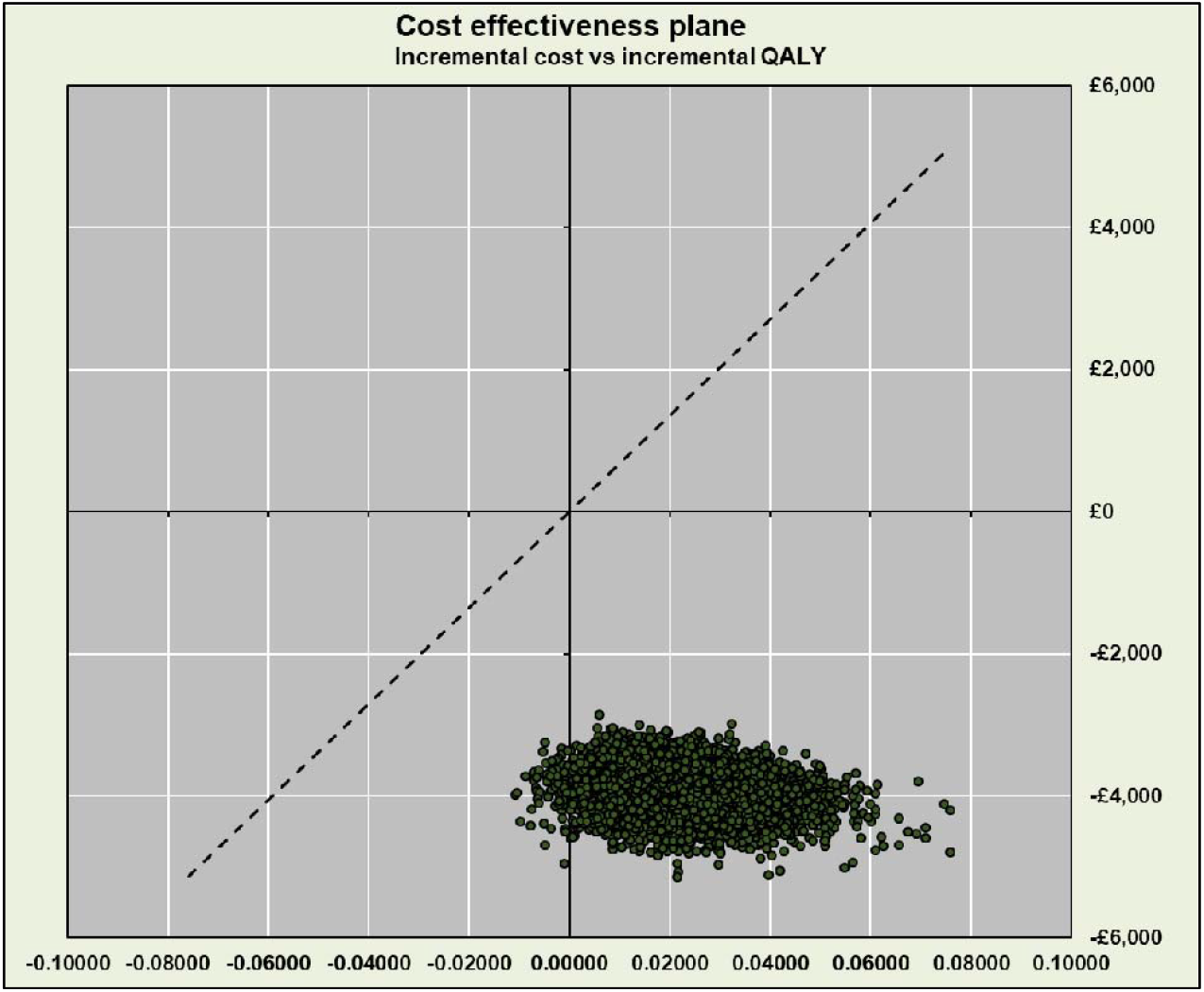
Scatter plot at £20,000 WTP threshold.

**Figure 3.**
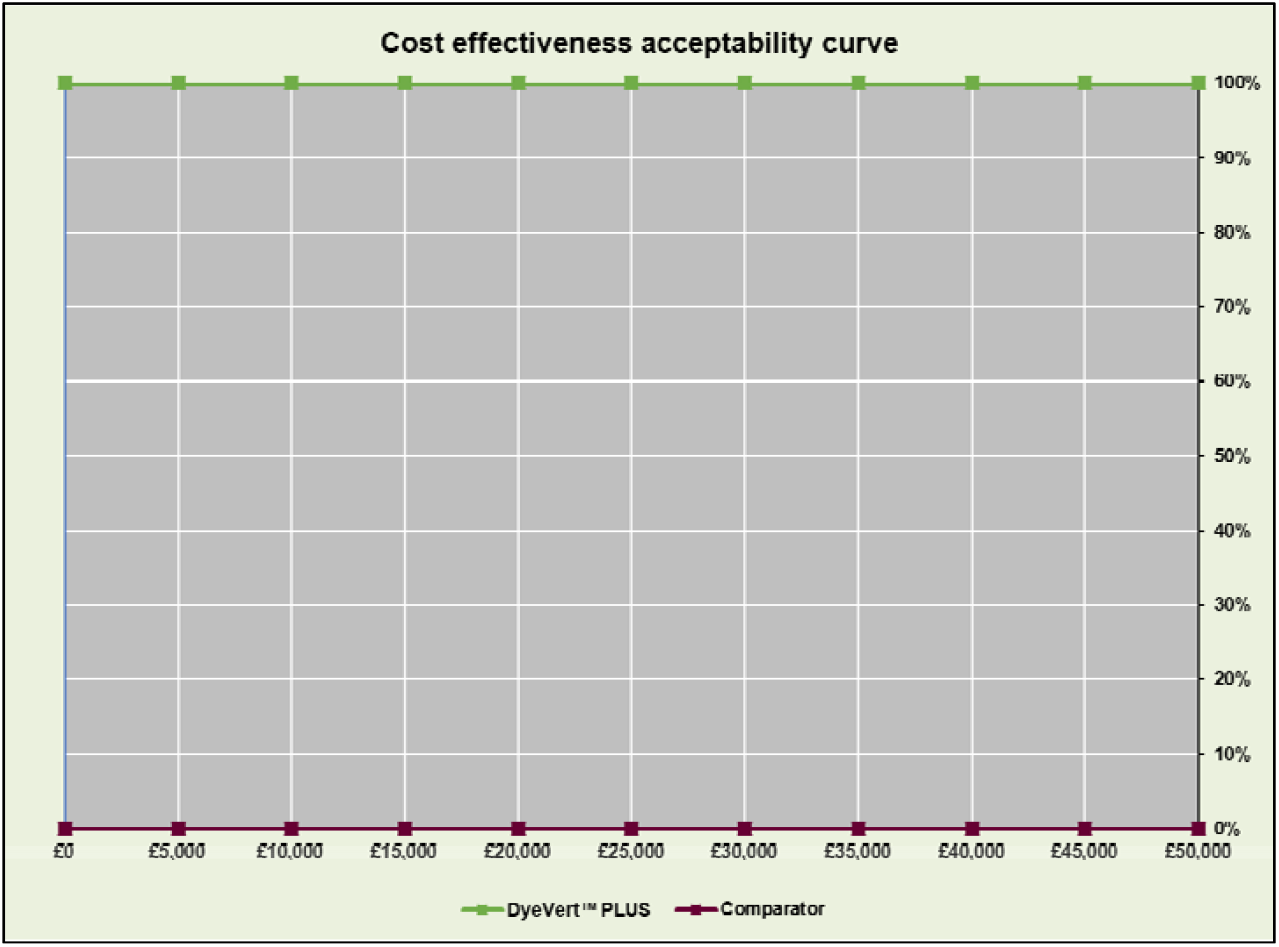
Cost-effectiveness acceptability curve at various WTP thresholds (£0-£50,000)

### Sensitivity analysis

A number of model parameters were also varied in sensitivity analysis. Multiple one-way sensitivity analyses were carried out to explore the impact of increasing/decreasing relevant parameter values by 25%. Figure 2 presents the impact of varying each parameter value by 25% on the incremental cost of the intervention, compared to current practice. Parameters are displayed in order, with those which have the greatest impact on incremental cost at the top and those which have the least impact at the end. In the base-case analysis, the incremental cost of the DyeVert™ PLUS EZ system was -£3,878. As seen in Figure 2 below, MI costs (in first and subsequent cycles) have the greatest impact on the incremental cost of the intervention (-/+ 12.3%). When this cost is decreased, cost savings of the intervention reduce and when this cost is increased, cost savings increase also. However, in both cases, DyeVert™ PLUS is still a cost saving strategy. All other parameters have minimal impact on the incremental costs.

Figure 5 and Figure 6 below shows the change in net monetary benefit (NMB) associated with varying each parameter by 25% and 100% in either direction. NMB is calculated as:

**Figure 4.**
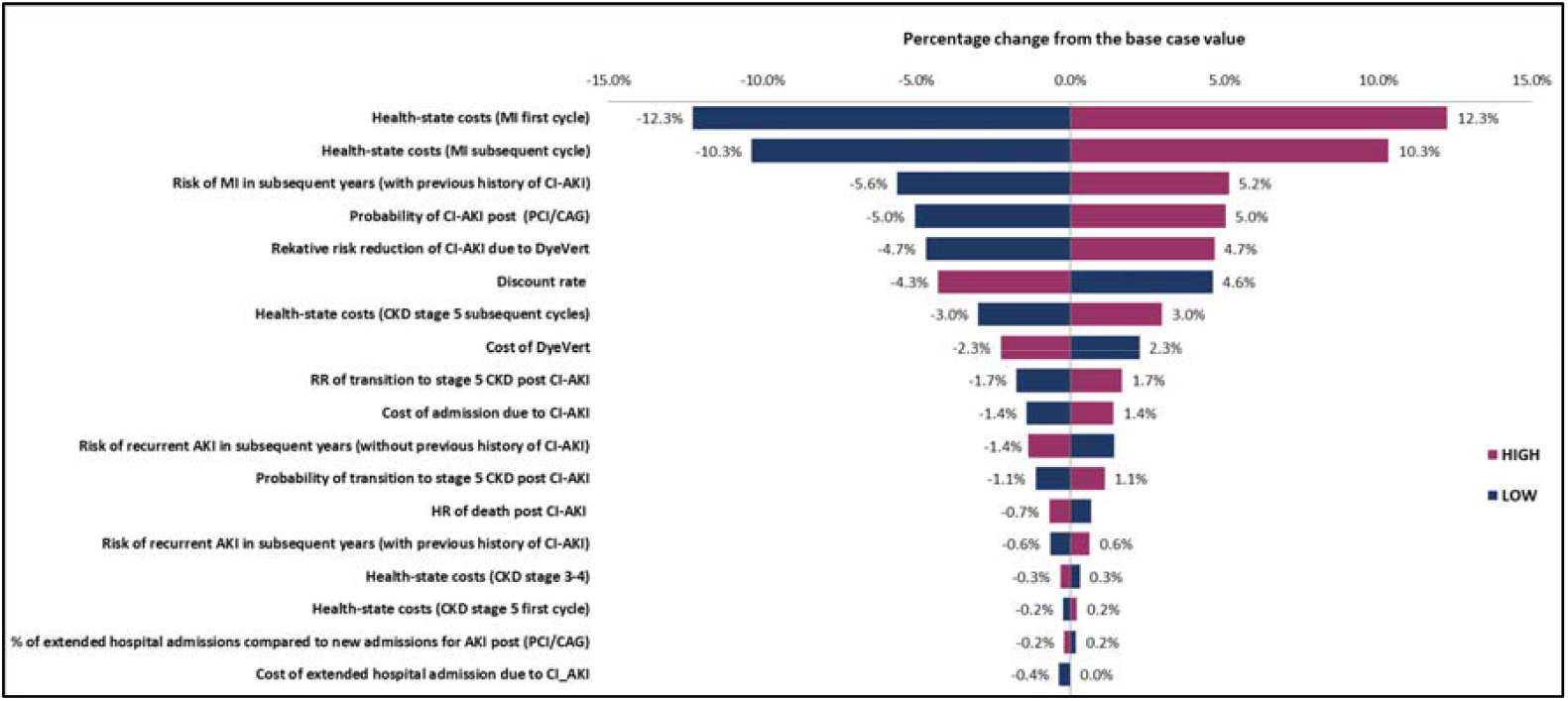
Tornado diagram showing the impact of changing the input parameters by ±25% on the estimated incremental cost.

**Figure 5.**
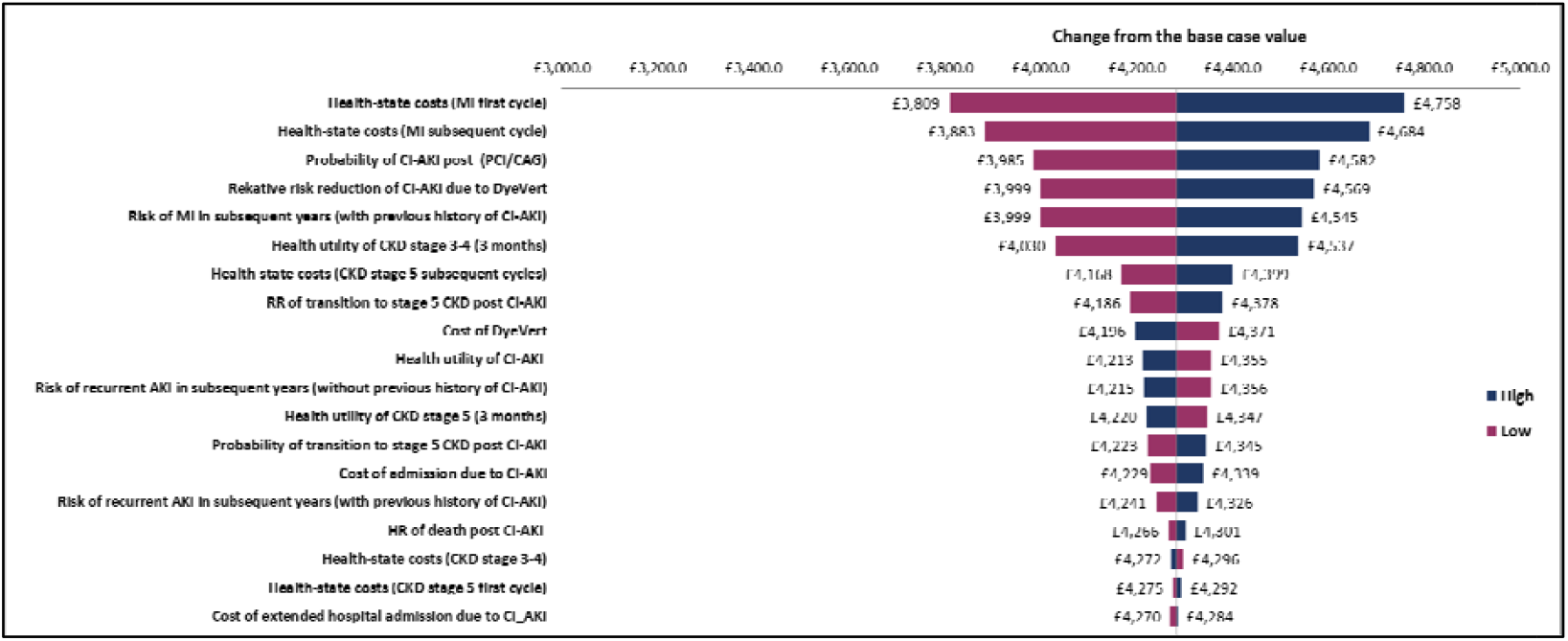
Impact of changing the input parameters by ±25% on the estimated NMB.

**Figure 6.**
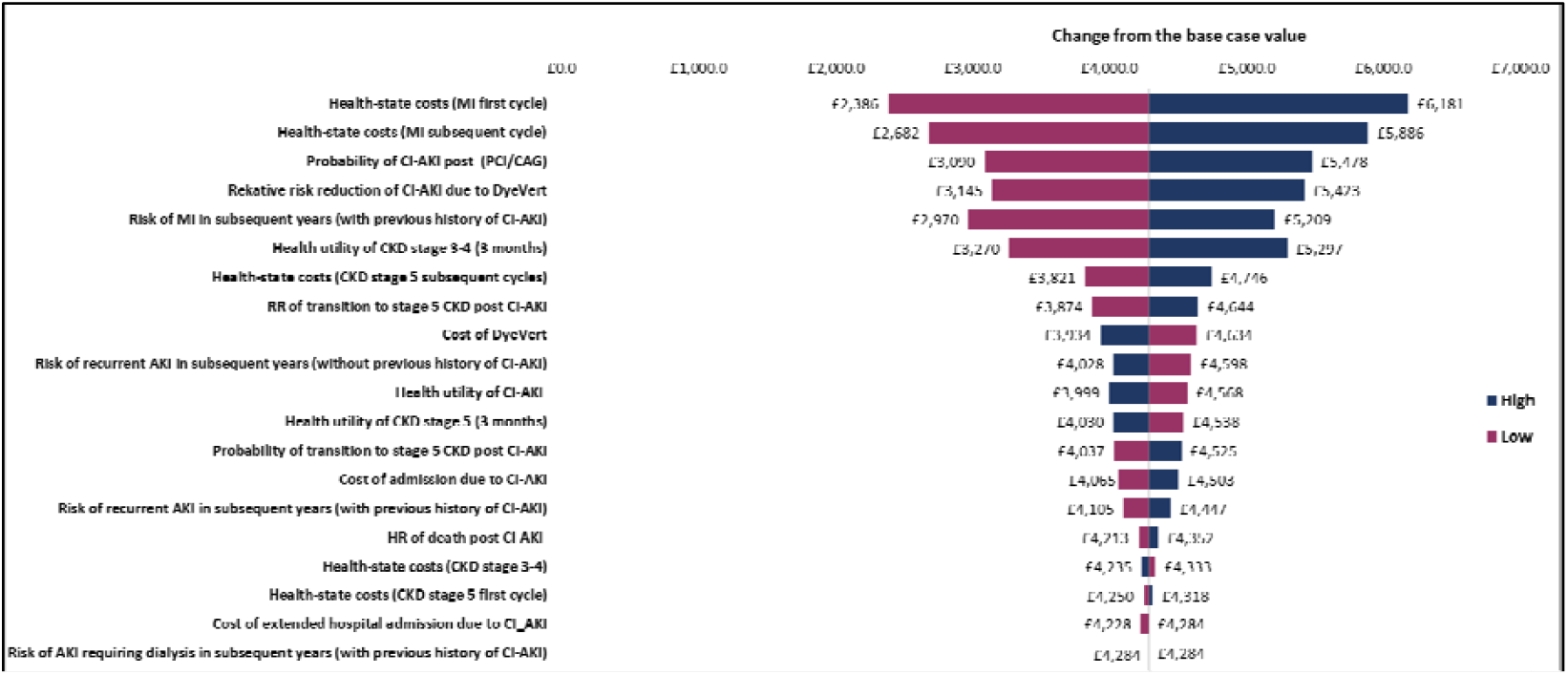
Impact of changing the input parameters by ±100% on the estimated NMB.

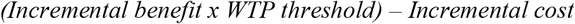

As in the previous analysis, varying the cost of the MI health state in the first and subsequent cycles is the greatest driver of change in overall results. However, regardless of the direction in which these costs are varied (± 25% and ±100%), the NMB of the intervention is still positive. Indeed, for all parameters included in this analysis, the NMB of the DyeVert™ PLUS EZ system remains positive, meaning that it is preferable to current practice from a health economic perspective.

## 5. Discussion

This study provides an insight into the potential cost savings that could be made by introducing the DyeVert™ PLUS EZ system into the UK health care system for use amongst patients with CKD stage 3-4 undergoing DAG and/or PCI. CI-AKI is one of the most common forms of AKI. The technology being evaluated is a device which acts by reducing the CMV, which has been shown to be linked to this specific form of the condition. The clinical efficacy of the technology has been demonstrated in previous clinical studies (13, 23, 36). Therefore, this innovative device has the potential to improve short-term health outcomes, and lead to cost savings and improved clinical outcomes in the longer-term. In order to assess costs and outcomes over the patient lifetime, a decision-analytic model was developed.

Results of the analysis presented here show the high likelihood that the DyeVert™ PLUS EZ system will be cost-effective over the lifetime of the patient. The probabilistic results were conclusive in that the device had a 100% probability of being cost-effective following 10,000 iterations of the model in a Monte Carlo simulation. Similarly, results of the deterministic sensitivity analyses indicated that the intervention would still be cost saving, and result in a positive net monetary benefit, in all scenarios assessed. Cost savings associated with introduction of the intervention are primarily related to the impact it has on the occurrence of AKI. Not only are there immediate cost savings associated with reducing the likelihood of experiencing an AKI, but there are also longer-term savings due to the relationship between AKI and development of CKD. See et al., 2018 (5) found that individuals who experienced AKI were at a heightened risk of experiencing new or progressive CKD. These long-term cost savings are captured in the model.

The number of studies that have been conducted to assess the relative cost-effectiveness of interventions to prevent onset of CI-AKI are limited. The National Clinical Guidelines Centre UK developed a Markov model in 2013, as part of their guidelines on AKI (24), to assess the cost-effectiveness of various different intravenous fluids for the prevention of the condition. Results of this study indicated that the most cost-effective strategy involved infusion with sodium chloride 0.9% and treatment with N-Acetylcysteine. Additionally, a study by Hiremath et al., 2018 (37) was identified which focussed on assessing the cost-effectiveness of utilising an iso-osmolar agent, iodixanol, compared to a low-osmolar contrast agent. Results of this study indicated that the iso-osmolar agent dominated the low-osmolar agent, i.e. was less costly and more effective over the patient’s lifetime. However, no economic evaluations focussing on the cost-effectiveness of interventions designed to divert an amount of the injected CMV were identified, which makes the study presented here unique.

There are limitations to the analysis presented. A number of assumptions were made in populating the model due to lack of appropriate data. Firstly, it was unknown how different the risk of developing CI-AKI would be depending on whether the patient had received DAG, PCI or a combination of both. Therefore, in the model it was assumed that this risk would be the same, regardless of which of the interventions was initially received. This may have the effect of over- or under-estimating the risk of developing this complication, depending on which initial intervention is received, and the direction of this effect is unknown. Secondly, there were no data to inform the utility value of patients with CKD stage 3-4 who have experienced a MI. Therefore, for the purpose of this analysis, it was assumed that the utility value of these patients would be same as the utility value of patients with CKD stage 3-4 who have not experienced this adverse event. Although the data used to inform the effectiveness of the DyeVert™ PLUS EZ system were derived from robust, published clinical evidence, the number of studies available to inform the clinical effectiveness of the device were limited. Ideally, when modelling the impact of a healthcare technology on clinical outcomes, multiple data sources would be available to verify the clinical efficacy data being used, and that was not the case for this analysis. Finally, although evidence exists on the relationship between AKI and long-term clinical outcomes (5, 12), information is limited which means there is also a degree of uncertainty around the relevant data included in the model. However an extreme sensitivity analysis was performed to address this limitation and the main input parameters were changed by ±100% but the conclusion remained stable. Additional minor assumptions were made in populating the model but none of these were likely to have a major impact on the final model results.

Despite the limitations highlighted above, a robust decision-analytic model was developed. The model was informed by clinical guidelines, published literature and expert clinical input, and any assumptions that were made in this analysis can be rectified by using more robust data in later studies, as a model now exists for re-analysis once additional information becomes available.

## 6. Conclusion

The economic analysis presented in this study has shown that introduction of the DyeVert™ PLUS EZ system has the potential to reduce costs for the UK health care service, and lead to improved quality of life and clinical outcomes for patients with CKD stage 3-4 undergoing angiographic procedures.

## Data Availability

All data are publicly available

## Data Availability Statement

The authors declare that all of the data supporting the findings of this study are available within the article (or the supplementary material of the article).

## Acknowledgment

We would like to show our gratitude to all team members who provided insight and expertise that greatly assisted the research.

## Compliance with Ethical Standards

### Author contributions

Mehdi Javanbakht and Atefeh Mashayekhi were responsible for developing and populating the economic model and drafting the first version of the manuscript. All authors provided inputs for the model, read, and approved the final draft of the manuscript.

